# Healthcare Worker Attendance During the Early Stages of the COVID-19 Pandemic: A Longitudinal Analysis of Daily Fingerprint-Verified Data from All Public-Sector Secondary and Tertiary Care Facilities in Bangladesh

**DOI:** 10.1101/2020.09.01.20186445

**Authors:** Duy Do, Malabika Sarker, Simiao Chen, Ali Lenjani, Pauli Tikka, Till Bärnighausen, Pascal Geldsetzer

**Affiliations:** Heidelberg Institute of Global Health, University of Heidelberg. Im Neuenheimer Feld 130.3 69120 Heidelberg, Germany; Division of Primary Care and Population Health, Department of Medicine, Stanford University. 1265 Welch Road, Stanford, CA 94305, USA; James P. Grant School of Public Health, BRAC University. 68 Shahid Tajuddin Ahmed Sharani, Mohakhali, Dhaka-1212, Bangladesh; Peking Union Medical College. 9 Dongdan 3rd Alley, Dong Dan, Dongcheng, Beijing, China; Harvard T.H. Chan School of Public Health, Harvard University. 677 Huntington Avenue, Boston, MA 02115, USA

**Author notes:** **Corresponding author:** Pascal Geldsetzer Division of Primary Care and Population Health Department of Medicine Stanford University 1265 Welch Road Stanford CA 94305, USA.

**Keywords:** COVID-19, healthcare workers, attendance, health system, global health

## Abstract

**Background:** The COVID-19 pandemic has overwhelmed hospitals in several areas in high-income countries. An effective response to this pandemic requires healthcare workers (HCWs) to be present at work, particularly in low- and middle-income countries (LMICs) where they are already in critically low supply. To inform whether and to what degree policymakers in Bangladesh, and LMICs more broadly, should expect a drop in HCW attendance as COVID-19 continues to spread, this study aims to determine how HCW attendance has changed during the early stages of the COVID-19 pandemic in Bangladesh.

**Methods:** This study analyzed daily fingerprint-verified attendance data from all 527 public-sector secondary and tertiary care facilities in Bangladesh to describe HCW attendance from January 26, 2019 to March 22, 2020, by cadre, hospital type, and geographic division. We then regressed HCW attendance onto fixed effects for day-of-week, month, and hospital, as well as indicators for each of three pandemic periods: a China-focused period (January 11, 2020 [first confirmed COVID-19 death in China] until January 29, 2020), international-spread period (January 30, 2020 [World Health Organization’s declaration of a global emergency] until March 6, 2020), and local-spread period (March 7, 2020 [first confirmed COVID-19 case in Bangladesh] until the end of the study period).

**Findings:** On average between January 26, 2019 and March 22, 2020, 34·1% of doctors, 64·6% of nurses, and 70·6% of other healthcare staff were present for their scheduled shift. HCWs’ attendance rate increased with time in 2019 among all cadres. Nurses’ attendance level dropped by 2·5% points (95% CI; –3·2% to –1·8%) and 3·5% points (95% CI; –4·5% to –2·5%) during the international-spread and the local-spread periods of the COVID-19 pandemic, relative to the China-focused period. Similarly, the attendance level of other healthcare staff declined by 0·3% points (95% CI; –0·8% to 0·2%) and 2·3% points (95% CI; –3·0% to –1·6%) during the international-spread and local-spread periods, respectively. Among doctors, however, the international-spread and local-spread periods were associated with a statistically significant increase in attendance by 3·7% points (95% CI; 2·5% to 4·8%) and 4·9% points (95% CI; 3·5% to 6·4%), respectively. The reduction in attendance levels across all HCWs during the local-spread period was much greater at large hospitals, where the majority of COVID-19 testing and treatment took place, than that at small hospitals.

**Conclusions:** After a year of significant improvements, HCWs’ attendance levels among nurses and other healthcare staff (who form the majority of Bangladesh’s healthcare workforce) have declined during the early stages of the COVID-19 pandemic. This finding may portend an even greater decrease in attendance if COVID-19 continues to spread in Bangladesh. Policymakers in Bangladesh and similar LMICs should undertake major efforts to achieve high attendance levels among HCWs, particularly nurses, such as by providing sufficient personal protective equipment as well as monetary and non-monetary incentives.

## I. Introduction

The novel coronavirus disease 2019 (COVID-19) was first reported in Wuhan, China in December 2019 [1]. Since then, COVID-19 has rapidly spread to more than 170 countries and has – according to confirmed cases – infected more than three and a half million individuals worldwide; 254,430 of whom have died from the infection as of May 5, 2020 [2,3]. COVID-19 has outstretched hospital capacity in high-resource settings in Europe and the United States [4,5]. If COVID-19 continues to spread in low- and middle-income countries (LMICs), these countries’ low-capacity secondary and tertiary care systems are likely to be quickly overwhelmed by the pandemic.

For LMICs to respond as effectively as possible to the pandemic will require that the health system is able to fully utilize its existing resources. Arguably the most essential health system resource is healthcare workers (HCWs). In addition to a critically low density of HCWs in most LMICs, many of these health systems also struggle with alarmingly low HCW attendance rates. In prior studies, HCW attendance rates have ranged from 52%-60% in Bangladesh, India, Indonesia, and Uganda to 75% in Peru and Kenya [6–9].

There is no empirical evidence so far on how HCW attendance rates at hospitals in LMICs will be affected by the COVID-19 pandemic. On the one hand, HCWs maybe more motivated to report to work because they feel compelled to contribute to the country’s pandemic response. On the other hand, HCW attendance may decline due to workers’ fear of infecting themselves and their families, especially given the lack of personal protective equipment (PPE) in many settings, or due to HCWs themselves falling ill with COVID-19. Previous studies suggested that HCWs were less likely to work during pandemics. The 2009 H1N1 influenza pandemic in Hong Kong was associated with an increase in the all-cause sickness absence rate by 57·7% for all healthcare cadres [10]. Similarly, the 1980–1981 pandemic of influenza A/Bangkok 79 was associated with a decline of 69·4% in HCW attendance in Thailand during the two-week peak of the pandemic relative to the same two weeks in the subsequent year when there was no pandemic [11].

Understanding whether and to what degree HCW attendance will change in LMICs during the COVID-19 epidemic could help policymakers take early action to increase attendance, such as ensuring a reliable and sufficient supply of PPE or providing monetary and non-monetary incentives to attend. This study leverages unique fingerprint-verified HCW attendance data collected daily at all public-sector secondary and tertiary care facilities in Bangladesh in 2019 and 2020 to i) describe the level of HCW attendance in Bangladesh and how this varies by healthcare cadres, hospital types, and geographic regions, and ii) to determine changes in HCW attendance during the early stages of the COVID-19 epidemic.

## II. Methods

### Study setting

Bangladesh is one of the world’s most populous countries, with a population estimation of more than 167 million people [12]. Similar to many other LMICs, Bangladesh is experiencing the double burden of communicable and noncommunicable disease as a result of rapid epidemiological and demographic changes, placing considerable strains on a generally understaffed and low-resourced healthcare system. Four sectors are responsible for the provision of medical care in Bangladesh – including the public sector, private sector, non-governmental organizations (NGOs), and donor agencies. The public sector is the main provider of medical care, with services ranging from curative, preventive, promotive, to rehabilitative care [13]. Public-sector health facilities offer medical care at various levels – including primary care at community clinics; secondary care at upazila health complexes (“sub-district hospitals” hereafter), district hospitals, and general hospitals; and tertiary care at medical college hospitals and specialized hospitals. Although public-sector health facilities serve a large proportion of the population, they are typically poorly equipped with medical supplies, are understaffed due to high vacancy and low attendance levels, and are unevenly distributed between rural and urban areas [9,13], which may hamper Bangladesh’s response to COVID-19.

As of May 4, 2020, the Institute of Epidemiology, Disease Control and Research (IEDCR) reported that the country had tested 87,694 individuals, 10,143 of whom were infected with the virus and 182 people died from infection [14] (see Figure 1 for the geographic distribution of COVID-19 confirmed cases). Testing and treatment were mostly centralized to the IEDCR in the capital city of Dhaka. The lack of testing has raised concerns about a potentially large number of undetected COVID-19 cases, which will expedite the spread of the virus, especially given the country’s high population density, a large proportion of communal living arrangements, and many migrants returning home from other heavily infected countries. As such, Bangladesh will likely face a surge in healthcare demand as a result of COVID-19, placing significant strains on its secondary and tertiary care system.

**Figure 1:**
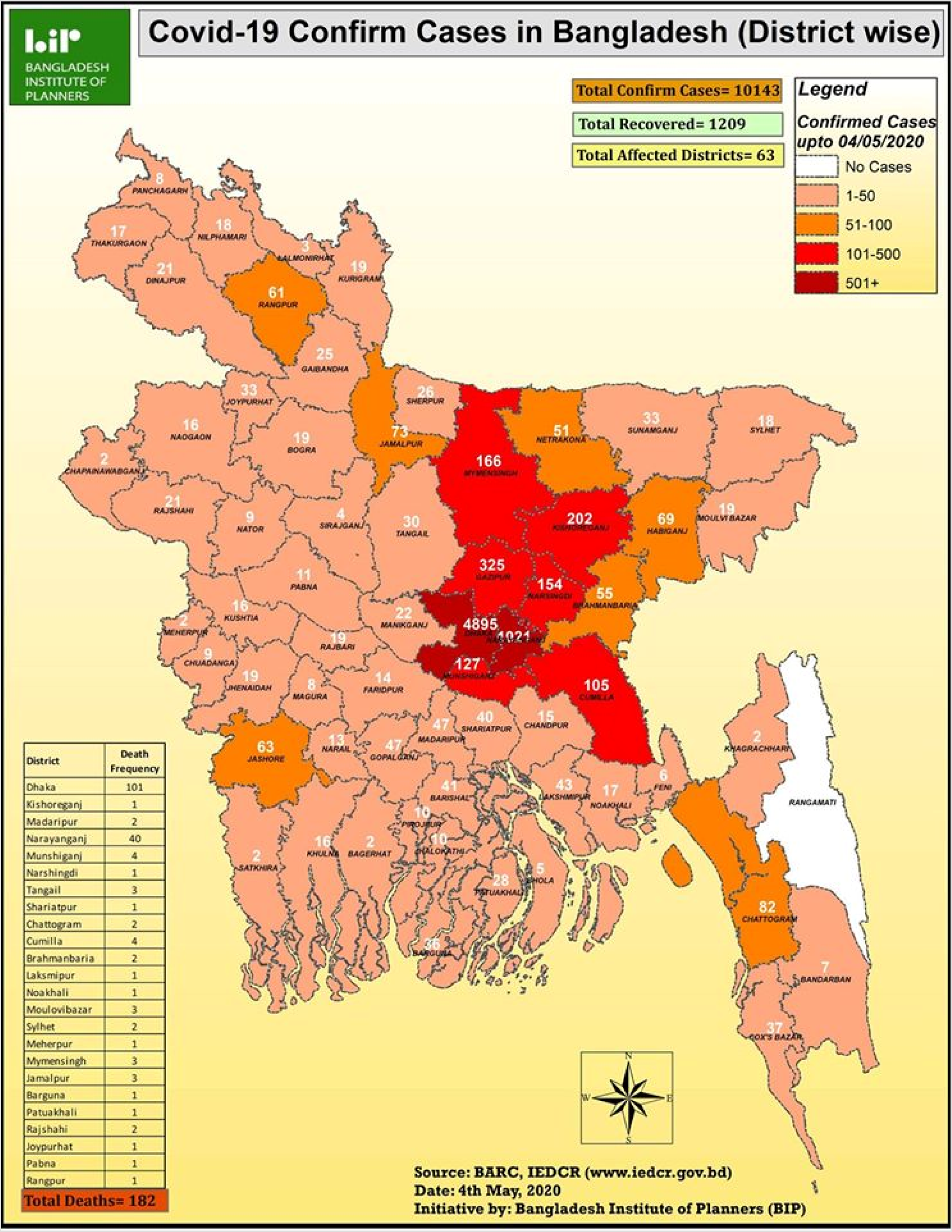
Geographic Distribution of Confirmed COVID-19 Cases in Bangladesh as of May 4, 2020.

Source: Bangladesh Institute of Planners.

### Data source

This study used attendance data from the Bangladesh’s Ministry of Health and Family Welfare collected daily at all public-sector secondary and tertiary medical facilities from January 26, 2019 to March 22, 2020. Attendance was verified using fingerprints rather than self-reports. All HCWs must scan their fingerprint using a machine sensor on arrival and departure. The date and time of arrival and departure is recorded directly in the central system, and aggregated staff attendance data is available to the public [15]. Data at the facility level included the hospital name; location; the daily number of doctors, nurses, and other staff (i.e. technicians, pharmacists, paramedics, and medical assistants) scheduled to work; and the daily number of each healthcare cadre present at work.

### Data analysis

Our analysis had three main steps. First, we calculated the daily percentage of workers present at work for each cadre as well as for all cadres at each facility. We eliminated administrative facilities and medical colleges that did not provide direct patient care (N = 46). The remaining 527 direct patient-care facilities comprised of all public-sector secondary and tertiary care facilities, which were categorized into hospitals/institutes – including medical college hospitals, hospitals with at least 100 beds, hospitals with fewer than 100 beds, general hospitals, district hospitals, and sub-district hospitals.

Second, using locally weighted robust regression (LOWESS), we described smoothed unadjusted trends in attendance levels of all HCWs as well as attendance of doctors, nurses, and other staff from January 26, 2019 to March 22, 2020.

Third, we investigated the association between the COVID-19 pandemic and HCW attendance from January 11, 2020 to March 22, 2020. We began with January 11, 2020 because this was the day on which the first COVID-19 death was confirmed in China. We ended the study on March 22, 2020 because the Bangladesh government ordered HCWs on this date to stop using fingerprint machines to record attendance due to fear that the practice of touching the fingerprint scanning device could spread SARS-CoV-2. The pandemic was divided into three periods based on the spread of COVID-19 internationally and within Bangladesh: the *China-focused period* (January 11^th^, 2020 [first confirmed COVID-19 death in China] until January 29^th^, 2020), the *international-spread period* (January 30^th^, 2020 [World Health Organization’s declaration of a global emergency] until March 6^th^, 2020), and the *local-spread period* (March 7^th^, 2020 [first confirmed COVID-19 case in Bangladesh] until the end of the study period). We used a multivariable ordinary least squares regression model to estimate absolute changes in the attendance rate for all healthcare cadres during the international-spread and the local-spread periods relative to the China-focused period (the reference category). In these regressions, we controlled for secular trends and time-invariant hospital characteristics by including a series of day-of-week fixed-effects, month fixed-effects, and hospital fixed-effects. Standard errors were adjusted for clustering by hospitals to account for possible heteroskedasticity and correlation in the error terms over time. We repeated the regression analysis by healthcare cadre (doctors, nurses, and other staff), hospital types (sub-district hospitals vs. other hospitals), and geographic divisions.

All analyses were conducted using Stata version 16·0 [16].

## III. Results

Table 1 provides descriptive statistics for the composition of hospital types and attendance rates in the full sample and across geographic divisions. Among 527 hospitals, a majority are sub-district hospitals (79·7%). Between January 26, 2019 and March 22, 2020, 104 HCWs on average per hospital were scheduled to work each day, 61·5% of whom were present at work. Attendance rates varied across cadres, ranging from 34·1% for doctors to 64·6% for nurses and 70·6% for other staff. We found little variation in hospital composition across divisions, except that the Dhaka division had more specialized hospitals and large hospitals with at least 100 beds than other divisions. Attendance rates of all HCWs were relatively similar across divisions, ranging from 59·1% in Mymensingh to 64·9% in Sylhet.

**Table 1:**
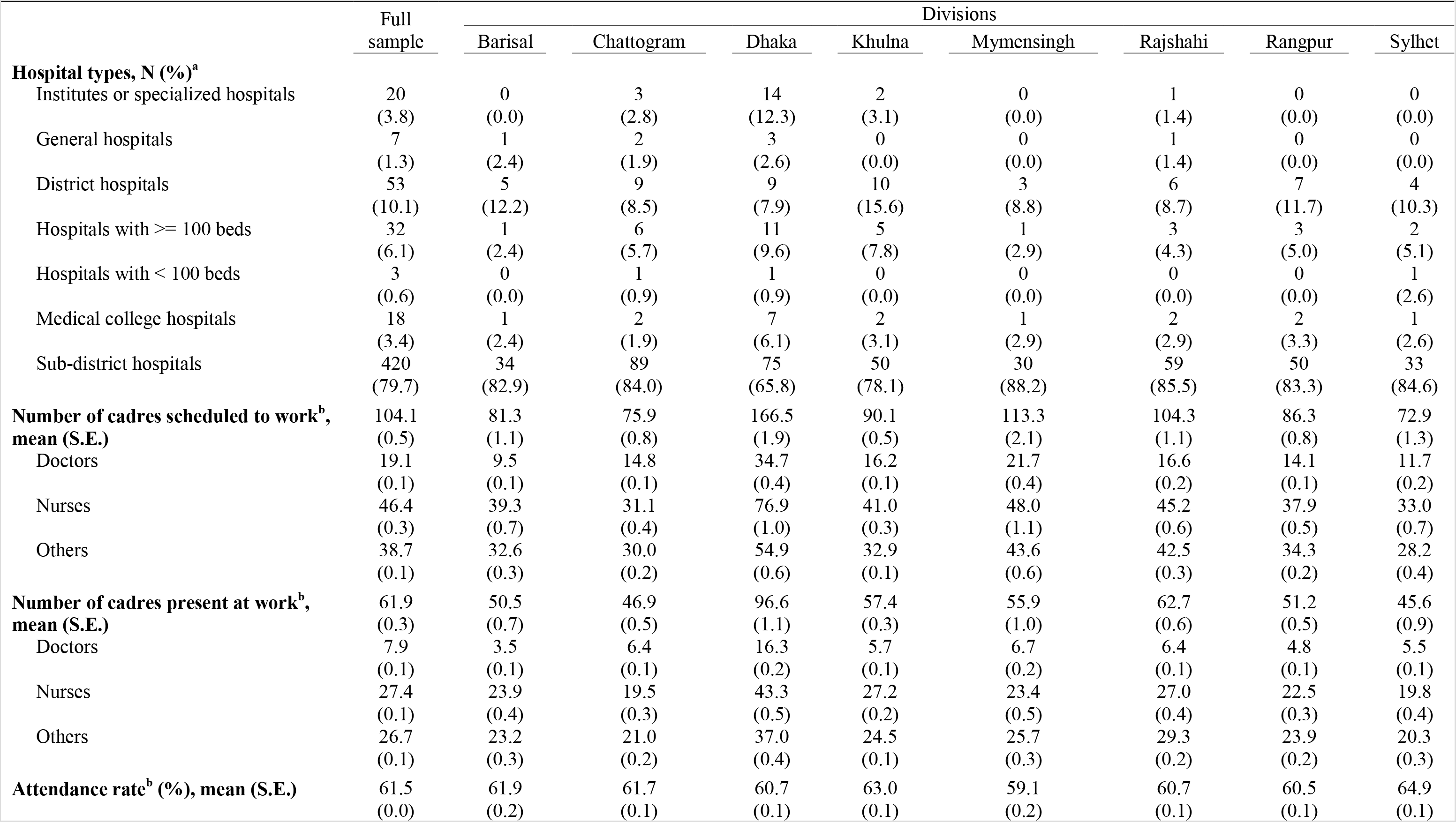
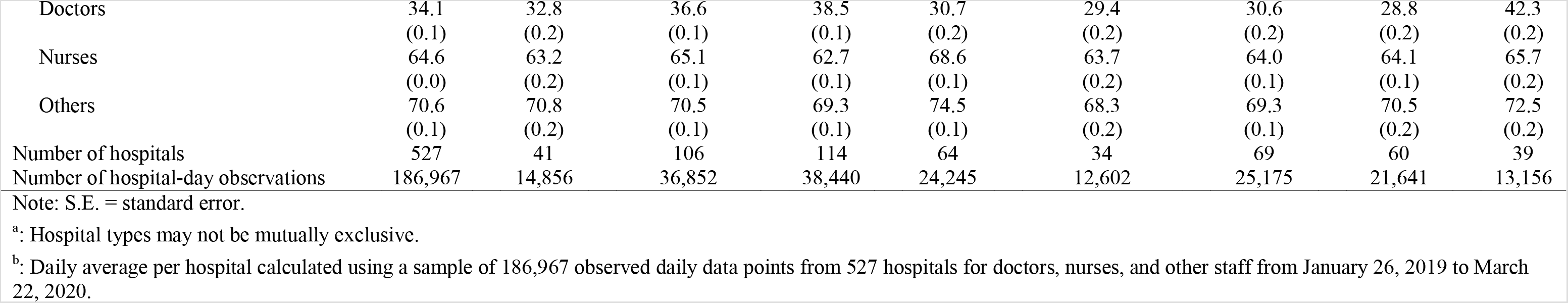
Descriptive Statistics of Hospital Types and Attendance Rates of Healthcare Workers at All Public-Sector Secondary and Tertiary Care Facilities in Bangladesh from January 26, 2019 to March 22, 2020.

|  | Full sample | Divisions |  |  |  |  |  |  |  |
| --- | --- | --- | --- | --- | --- | --- | --- | --- | --- |
|  |  | Barisal | Chattogram | Dhaka | Khulna | Mymensingh | Rajshahi | Rangpur | Sylhet |
| **Hospital types, N (%)<sup>a</sup>** |  |  |  |  |  |  |  |  |  |
| Institutes or specialized hospitals | 20<br>(3.8) | 0<br>(0.0) | 3<br>(2.8) | 14<br>(12.3) | 2<br>(3.1) | 0<br>(0.0) | 1<br>(1.4) | 0<br>(0.0) | 0<br>(0.0) |
| General hospitals | 7<br>(1.3) | 1<br>(2.4) | 2<br>(1.9) | 3<br>(2.6) | 0<br>(0.0) | 0<br>(0.0) | 1<br>(1.4) | 0<br>(0.0) | 0<br>(0.0) |
| District hospitals | 53<br>(10.1) | 5<br>(12.2) | 9<br>(8.5) | 9<br>(7.9) | 10<br>(15.6) | 3<br>(8.8) | 6<br>(8.7) | 7<br>(11.7) | 4<br>(10.3) |
| Hospitals with ≥ 100 beds | 32<br>(6.1) | 1<br>(2.4) | 6<br>(5.7) | 11<br>(9.6) | 5<br>(7.8) | 1<br>(2.9) | 3<br>(4.3) | 3<br>(5.0) | 2<br>(5.1) |
| Hospitals with < 100 beds | 3<br>(0.6) | 0<br>(0.0) | 1<br>(0.9) | 1<br>(0.9) | 0<br>(0.0) | 0<br>(0.0) | 0<br>(0.0) | 0<br>(0.0) | 1<br>(2.6) |
| Medical college hospitals | 18<br>(3.4) | 1<br>(2.4) | 2<br>(1.9) | 7<br>(6.1) | 2<br>(3.1) | 1<br>(2.9) | 2<br>(2.9) | 2<br>(3.3) | 1<br>(2.6) |
| Sub-district hospitals | 420<br>(79.7) | 34<br>(82.9) | 89<br>(84.0) | 75<br>(65.8) | 50<br>(78.1) | 30<br>(88.2) | 59<br>(85.5) | 50<br>(83.3) | 33<br>(84.6) |
| **Number of cadres scheduled to work<sup>b</sup>, mean (S.E.)** | 104.1<br>(0.5) | 81.3<br>(1.1) | 75.9<br>(0.8) | 166.5<br>(1.9) | 90.1<br>(0.5) | 113.3<br>(2.1) | 104.3<br>(1.1) | 86.3<br>(0.8) | 72.9<br>(1.3) |
| Doctors | 19.1<br>(0.1) | 9.5<br>(0.1) | 14.8<br>(0.1) | 34.7<br>(0.4) | 16.2<br>(0.1) | 21.7<br>(0.4) | 16.6<br>(0.2) | 14.1<br>(0.1) | 11.7<br>(0.2) |
| Nurses | 46.4<br>(0.3) | 39.3<br>(0.7) | 31.1<br>(0.4) | 76.9<br>(1.0) | 41.0<br>(0.3) | 48.0<br>(1.1) | 45.2<br>(0.6) | 37.9<br>(0.5) | 33.0<br>(0.7) |
| Others | 38.7<br>(0.1) | 32.6<br>(0.3) | 30.0<br>(0.2) | 54.9<br>(0.6) | 32.9<br>(0.1) | 43.6<br>(0.6) | 42.5<br>(0.3) | 34.3<br>(0.2) | 28.2<br>(0.4) |
| **Number of cadres present at work<sup>b</sup>, mean (S.E.)** | 61.9<br>(0.3) | 50.5<br>(0.7) | 46.9<br>(0.5) | 96.6<br>(1.1) | 57.4<br>(0.3) | 55.9<br>(1.0) | 62.7<br>(0.6) | 51.2<br>(0.5) | 45.6<br>(0.9) |
| Doctors | 7.9<br>(0.1) | 3.5<br>(0.1) | 6.4<br>(0.1) | 16.3<br>(0.2) | 5.7<br>(0.1) | 6.7<br>(0.2) | 6.4<br>(0.1) | 4.8<br>(0.1) | 5.5<br>(0.1) |
| Nurses | 27.4<br>(0.1) | 23.9<br>(0.4) | 19.5<br>(0.3) | 43.3<br>(0.5) | 27.2<br>(0.2) | 23.4<br>(0.5) | 27.0<br>(0.4) | 22.5<br>(0.3) | 19.8<br>(0.4) |
| Others | 26.7<br>(0.1) | 23.2<br>(0.3) | 21.0<br>(0.2) | 37.0<br>(0.4) | 24.5<br>(0.1) | 25.7<br>(0.3) | 29.3<br>(0.2) | 23.9<br>(0.2) | 20.3<br>(0.3) |
| **Attendance rate<sup>b</sup> (%), mean (S.E.)** | 61.5<br>(0.0) | 61.9<br>(0.2) | 61.7<br>(0.1) | 60.7<br>(0.1) | 63.0<br>(0.1) | 59.1<br>(0.2) | 60.7<br>(0.1) | 60.5<br>(0.1) | 64.9<br>(0.1) |
| Doctors | 34.1<br>(0.1) | 32.8<br>(0.2) | 36.6<br>(0.1) | 38.5<br>(0.1) | 30.7<br>(0.2) | 29.4<br>(0.2) | 30.6<br>(0.2) | 28.8<br>(0.2) | 42.3<br>(0.2) |
| Nurses | 64.6<br>(0.0) | 63.2<br>(0.2) | 65.1<br>(0.1) | 62.7<br>(0.1) | 68.6<br>(0.1) | 63.7<br>(0.2) | 64.0<br>(0.1) | 64.1<br>(0.1) | 65.7<br>(0.2) |
| Others | 70.6<br>(0.1) | 70.8<br>(0.2) | 70.5<br>(0.1) | 69.3<br>(0.1) | 74.5<br>(0.1) | 68.3<br>(0.2) | 69.3<br>(0.1) | 70.5<br>(0.2) | 72.5<br>(0.2) |
| Number of hospitals | 527 | 41 | 106 | 114 | 64 | 34 | 69 | 60 | 39 |
| Number of hospital-day observations | 186,967 | 14,856 | 36,852 | 38,440 | 24,245 | 12,602 | 25,175 | 21,641 | 13,156 |

Note: S.E. = standard error.
<sup>a</sup>: Hospital types may not be mutually exclusive.
<sup>b</sup>: Daily average per hospital calculated using a sample of 186,967 observed daily data points from 527 hospitals for doctors, nurses, and other staff from January 26, 2019 to March 22, 2020.

Figure 2 presents the smoothed unadjusted trends in attendance for all healthcare cadres at all secondary and tertiary care facilities from 2019 to 2020 (see Appendix Figure 1 for the unsmoothed unadjusted trend in weekly attendance). Overall, the average attendance rate was low, but increased gradually over the course of the study period. In January 2019, 45·5% (95% CI; 44·6% to 46·3%) of all HCWs were present at work, while this estimate increased to 63·1% (95% CI; 62·8% to 63·3%) by December 2019. HCW attendance levels continued to increase during the China-focused period and the international-spread period. However, attendance started to level off and then declined after the occurrence of the first confirmed COVID-19 case in Bangladesh on March 7, 2020.

**Figure 2:**
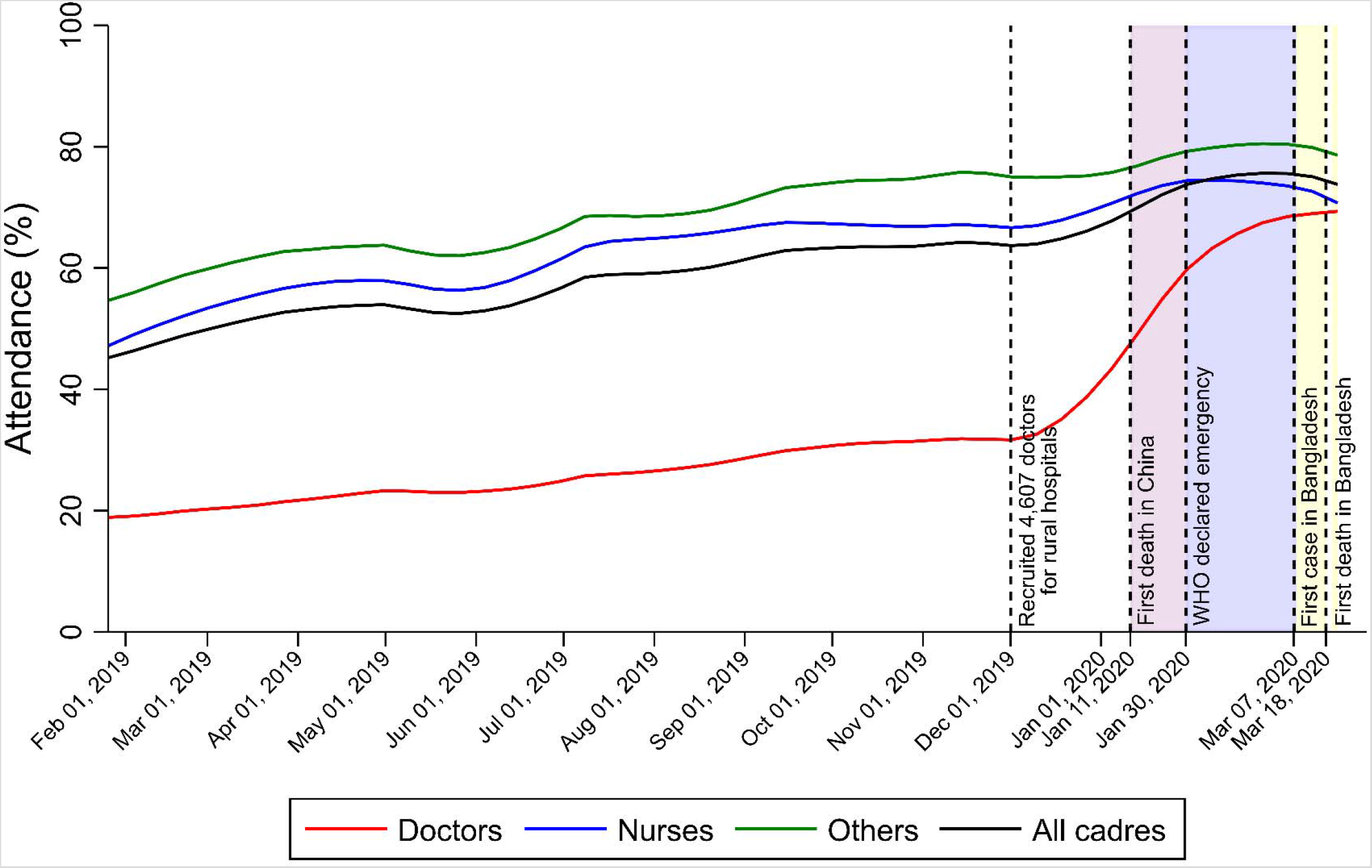
LOWESS-Estimated (Unadjusted) Trends in Attendance Rates Among Doctors, Nurses, and Other Staff at All Public-Sector Secondary and Tertiary Care Facilities in Bangladesh, by Cadre.

Further investigation revealed different patterns across healthcare cadres. Among doctors, the attendance rate was significantly lower than those of nurses and other staff in 2019. Doctors’ attendance rates increased significantly in December 2019 following the recruitment of 4,607 doctors at sub-district hospitals in rural areas, from 31·1% (95% CI; 30·8% to 31·5%) in November 2019 to 42·2% (95% CI; 41·3% to 43·1%) during the first week of January 2020. Doctors’ attendance further increased to 57·1% (95% CI; 56·6% to 57·6%) during the China-focused period, and then continued growing to 65·7% (95% CI; 65·3% to 66·0%) and 70.4% (95% CI; 69·9% to 70·9%) during the international-spread and the local-spread periods, respectively. The attendance levels of nurses and other staff followed a different pattern. In 2019, the attendance levels of nurses and other staff were much higher than that of doctors (62·0% [95% CI; 61·9 to 62·1%] and 68·2%% [95% CI; 68·1% to 68·3%] for nurses and other staff, respectively, versus 26·5% [95% CI; 26·4% to 26·6%] for doctors). Attendance levels of nurses and other staff increased during the China-focused period and then started to decline after the occurrence of the first COVID-19 case in Bangladesh.

Trend analyses for each of the eight geographic divisions (Figure 3) and by hospital types (Figure 4) suggested similar patterns as observed in Figure 2. Most divisions experienced a decrease in HCW attendance levels during the local-spread period. Figure 4 shows that doctors’ low attendance rate in 2019 and the significant increase in their attendance at the end of 2019 were mainly driven by those working at small sub-district hospitals (Panel B), rather than at large hospitals (Panel A). In addition, the reduction in attendance levels across all HCWs during the local-spread period was much greater at large hospitals than that at small sub-district hospitals.

**Figure 3:**
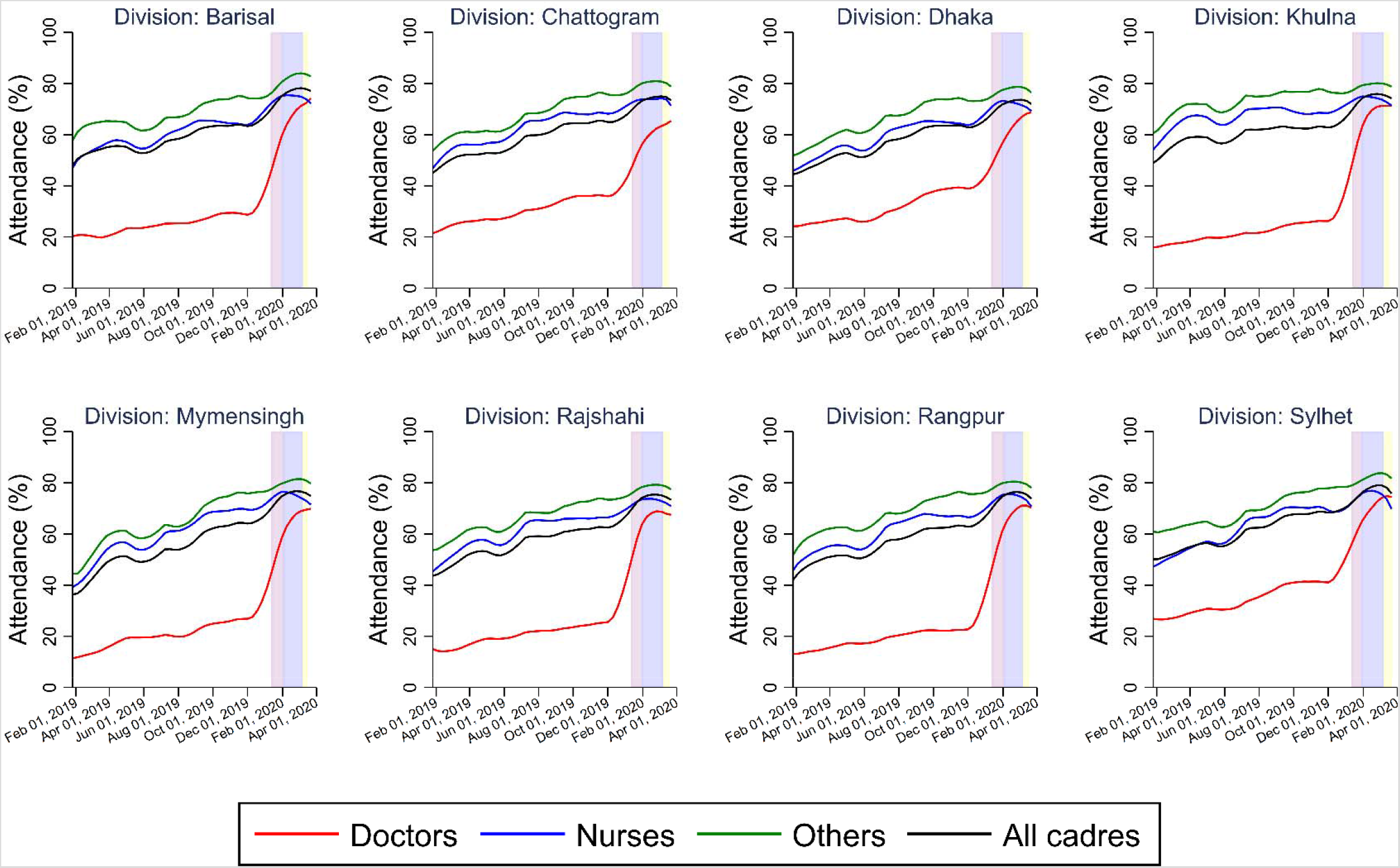
LOWESS-Estimated (Unadjusted) Trends in Attendance Rates Among Doctors, Nurses, and Other Staff at All Public-Sector Secondary and Tertiary Care Facilities in Bangladesh, by Cadre and Division.

**Figure 4:**
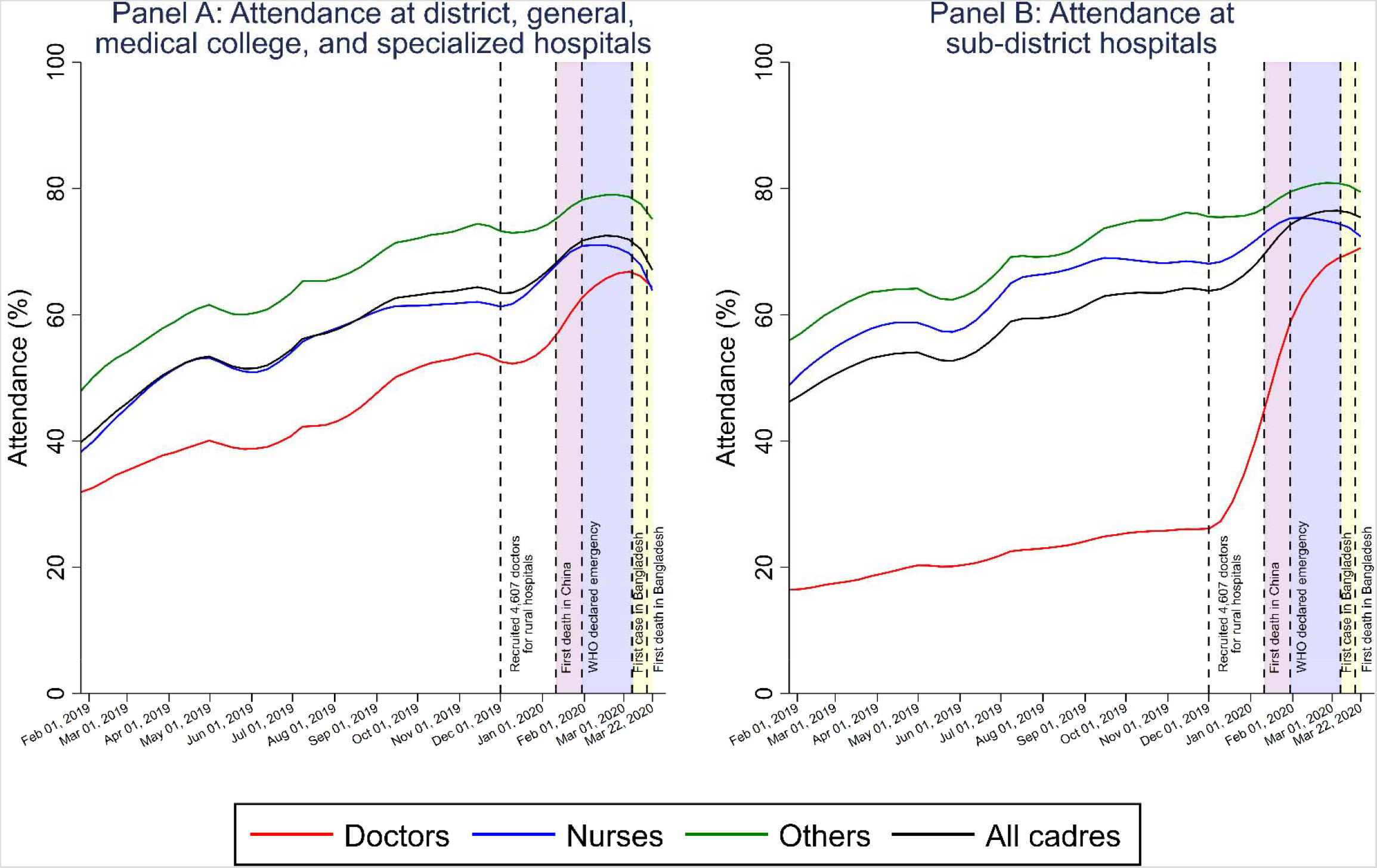
LOWESS-Estimated (Unadjusted) Trends in Attendance Rates Among Doctors, Nurses, and Other Staff at All Public-Sector Secondary and Tertiary Care Facilities in Bangladesh, by Hospital Type.

Table 2 quantifies the association between the early stages of the COVID-19 pandemic and changes in HCW attendance. In column (1), the international-spread and the local-spread periods were associated with an absolute decline in HCW attendance levels by 0·5% points (95% CI; –0·9% to –0·04%) and 1·7% points (95% CI; –2·3% to –1·1%), respectively. The association between the pandemic periods and HCW attendance was greater at large hospitals (column 2) compared to that at sub-district hospitals (column 3). The results were fairly consistent across geographic divisions (columns 4 to 11). HCW attendance rates generally declined in most divisions during the local-spread period compared to the China-focused period.

**Table 2:**
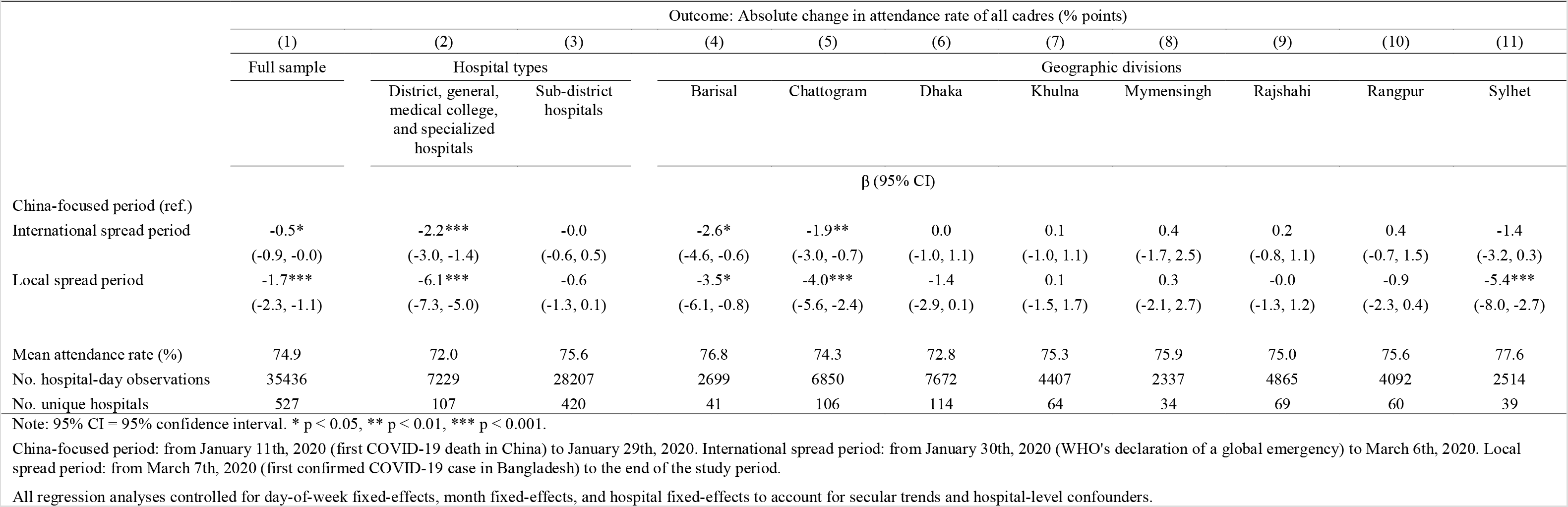
Linear Regression Estimates of Attendance Rates of All Healthcare Cadres on COVID-19 Pandemic Periods, by Hospital Type and Division.

The association between the pandemic periods and attendance varied across healthcare cadres (Table 3). Using the full sample of 527 hospitals, results in column (1) show that the international-spread and local-spread periods were associated with a statistically significant increase in attendance levels for doctors by 3·7% points (95% CI; 2·5% to 4·8%) and 4·9% points (95% CI; 3·5% to 6·4%), respectively, compared to that during the China-focused period (Panel A). Additional analyses stratified by hospital types indicated that doctors’ attendance at larger hospitals slightly increased by 0·5% points (95% CI; –0·8% to 1·9%) during the international-spread period and then declined by 0·6% points (95% CI; –2·5% to 1·3%) during the local-spread period (column 2). In contrast, doctors’ attendance rate increased significantly during the early stages of the pandemic at sub-district hospitals (column 3). Attendance levels of nurses and other staff immediately declined during the international-spread and local-spread periods (Panels B and C of Table 3). Nurses’ attendance level dropped by 2·5% points (95% CI; –3·2% to –1·8%) and 3·5% points (95% CI; –4·5% to –2·5%) during the international-spread and the local-spread periods, relative to the China-focused period. Similarly, the attendance level of other staff declined by 0·3% points (95% CI; –0·8% to 0·2%) and 2·3% points (95% CI; –3·0% to –1·6%) during the international-spread and local-spread periods, respectively. The association between the pandemic periods and attendance levels among nurses and other staff was driven more by declining attendance at large hospitals (column 2), rather than at sub-district hospitals (column 3).

**Table 3:** Linear Regression Estimates of Attendance Rates of Doctors, Nurses, and Other Staff on COVID-19 Pandemic Periods, by Hospital Type and Division.

|  | Outcome: Absolute change in attendance rates (% points) |  |  |  |  |  |  |  |  |  |  |
| --- | --- | --- | --- | --- | --- | --- | --- | --- | --- | --- | --- |
|  | (1) | (2) | (3) | (4) | (5) | (6) | (7) | (8) | (9) | (10) | (11) |
|  | Full sample | Hospital types |  | Geographic divisions |  |  |  |  |  |  |  |
|  |  | District, general, medical college, and specialized hospitals | Sub-district hospitals | Barisal | Chattogram | Dhaka | Khulna | Mymensingh | Rajshahi | Rangpur | Sylhet |
| β (95% CI) |  |  |  |  |  |  |  |  |  |  |  |
| **Panel A: Doctors** |  |  |  |  |  |  |  |  |  |  |  |
| China-focused period (ref.) |  |  |  |  |  |  |  |  |  |  |  |
| International spread period | 3.7***<br>(2.5, 4.8) | 0.5<br>(-0.8, 1.9) | 4.4***<br>(3.0, 5.8) | 1.3<br>(-3.3, 5.8) | 2.8*<br>(0.3, 5.2) | 5.0***<br>(2.5, 7.5) | 5.3**<br>(2.1, 8.5) | 3.9<br>(-2.9, 10.6) | 1.7<br>(-0.8, 4.3) | 7.1***<br>(3.7, 10.4) | -1.7<br>(-7.1, 3.7) |
| Local spread period | 4.9***<br>(3.5, 6.4) | -0.6<br>(-2.5, 1.3) | 6.3***<br>(4.6, 8.0) | 6.3*<br>(0.3, 12.3) | 5.0***<br>(2.2, 7.9) | 4.4**<br>(1.2, 7.6) | 8.0***<br>(4.2, 11.8) | 5.5<br>(-1.6, 12.5) | 3.5*<br>(0.4, 6.6) | 6.9**<br>(2.7, 11.2) | -2.7<br>(-10.0, 4.6) |
| Mean attendance rate (%) | 64.5 | 65.0 | 64.3 | 65.6 | 60.0 | 61.7 | 68.0 | 64.1 | 67.0 | 66.4 | 69.6 |
| **Panel B: Nurses** |  |  |  |  |  |  |  |  |  |  |  |
| China-focused period (ref.) |  |  |  |  |  |  |  |  |  |  |  |
| International spread period | -2.5***<br>(-3.2, -1.8) | -3.6***<br>(-4.7, -2.5) | -2.2***<br>(-3.1, -1.4) | -6.6***<br>(-9.8, -3.4) | -3.7***<br>(-5.6, -1.7) | -2.7***<br>(-4.1, -1.3) | -3.4***<br>(-4.9, -1.8) | -0.3<br>(-3.2, 2.5) | -0.6<br>(-2.2, 0.9) | -1.0<br>(-2.7, 0.7) | -1.5<br>(-5.1, 2.0) |
| Local spread period | -3.5***<br>(-4.5, -2.5) | -8.3***<br>(-9.8, -6.8) | -2.3***<br>(-3.5, -1.2) | -8.3***<br>(-12.8, -3.9) | -7.4***<br>(-10.0, -4.8) | -2.8*<br>(-4.9, -0.7) | -2.6*<br>(-4.7, -0.5) | 2.8<br>(-1.1, 6.6) | 0.8<br>(-1.3, 3.0) | -3.0*<br>(-5.4, -0.6) | -7.4***<br>(-11.4, -3.4) |
| Mean attendance rate (%) | 74.1 | 70.5 | 75.1 | 75.2 | 74.1 | 72.6 | 74.5 | 75.7 | 73.4 | 75.0 | 75.9 |
| **Panel C: Other staff** |  |  |  |  |  |  |  |  |  |  |  |
| China-focused period (ref.) |  |  |  |  |  |  |  |  |  |  |  |
| International spread period | -0.3<br>(-0.8, 0.2) | -0.8<br>(-1.8, 0.2) | -0.2<br>(-0.8, 0.5) | 0.1<br>(-2.2, 2.3) | -2.2**<br>(-3.6, -0.7) | 0.4<br>(-0.8, 1.6) | 0.6<br>(-0.8, 1.9) | -0.0<br>(-1.7, 1.7) | 0.4<br>(-0.7, 1.5) | -0.5<br>(-1.7, 0.7) | -1.1<br>(-3.3, 1.0) |
| Local spread period | -2.3***<br>(-3.0, -1.6) | -4.2***<br>(-5.7, -2.7) | -1.9***<br>(-2.7, -1.1) | -1.9<br>(-4.6, 0.9) | -4.9***<br>(-6.9, -3.0) | -1.8*<br>(-3.4, -0.2) | -0.8<br>(-2.7, 1.1) | -2.5*<br>(-4.4, -0.6) | -1.3<br>(-2.8, 0.2) | -1.5<br>(-3.0, 0.1) | -4.3**<br>(-7.2, -1.5) |
| Mean attendance rate (%) | 80.0 | 78.6 | 80.3 | 82.3 | 80.7 | 78.1 | 79.9 | 80.6 | 79.0 | 80.2 | 82.4 |
| No. hospital-day observations | 35436 | 7229 | 28207 | 2699 | 6850 | 7672 | 4407 | 2337 | 4865 | 4092 | 2514 |
| No. unique hospitals | 527 | 107 | 420 | 41 | 106 | 114 | 64 | 34 | 69 | 60 | 39 |

Note: 95% CI = 95% confidence interval. \* $p < 0.05$ , \*\* $p < 0.01$ , \*\*\* $p < 0.001$ .

## IV. Discussion

To our knowledge, this study is the first to investigate how HCW attendance in LMICs is changing during the early stages of the COVID-19 pandemic. Using daily fingerprint-verified attendance data from all public-sector secondary and tertiary care facilities in Bangladesh in 2019–2020, we documented a modest but significant decline in HCW attendance during the early stages of the COVID-19 pandemic, raising concerns about further declines in HCW attendance as COVID-19 continues to spread in Bangladesh and other LMICs. Our main findings are fourfold. First, the average attendance rate across all HCWs and medical facilities was low in 2019 but increased gradually over the course of the study period. Second, HCW attendance rates continued to increase in early 2020 during the China-focused period but have declined since the WHO’s declaration of a global emergency and the first confirmed case of COVID-19 in Bangladesh. Third, we observed important differences between doctors and other cadres in how the COVID-19 pandemic was associated with attendance, with attendance levels of nurses and other staff decreasing during the early stages of the COVID-19 pandemic, while doctors’ attendance level increased. Finally, the decline in HCW attendance during the early stages of the COVID-19 pandemic was greater at large hospitals where COVID-19 testing and treatment primarily took place compared to that at sub-district hospitals, suggesting that HCWs’ perception of increased risks of infection may influence their attendance.

The association between the early stages of the COVID-19 pandemic and HCW attendance documented in this study was lower compared to findings from previous pandemics. Using administrative data from Hong Kong, Ip and colleague [10] demonstrated a significant increase in all-cause absenteeism among HCWs at public medical facilities during the 2009 H1N1 influenza pandemic. Following the occurrence of the first untraceable local case of H1N1 influenza in Hong Kong, all-cause sickness absence rate increased from that in the baseline non-epidemic period by a relative amount of 57·7% for all healthcare cadres, 142·1% for medical staff, 32·7% for nursing staff, 94·5% for allied-health staff, and 77·6% for supporting staff. A second study by Hammond and Cheang [11] found that the 1980–1981 pandemic of influenza A/Bangkok 79 was associated with a relative decrease of 69·4% in the attendance rate at a teaching hospital in Thailand during the two-week peak of the pandemic compared to the same two weeks in the subsequent year when there was no pandemic. In comparison, our study found that the occurrence of the first confirmed COVID-19 case in Bangladesh was associated with an absolute decline in attendance levels by 1·7% points for all healthcare cadres, 3·5% points for nurses, and 2·3% points for other staff compared to those in the baseline period (when the epidemic was concentrated in China), which equates to a relative decline of 2·2%, 4·7%, and 2·9%, respectively. The main reason for which the decreases we observed in HCW attendance were lower than those in studies of other pandemics likely is that we were only able to observe trends during the early stages of the COVID-19 pandemic. Our findings suggest that if COVID-19 continues to spread in Bangladesh, the trends in declining HCW attendance that we observed during the early stages of the pandemic will probably continue or even accelerate.

The greatest declines in attendance levels during the early stages of the COVID-19 pandemic in Bangladesh were among nurses, followed by other staff. In contrast, doctors’ attendance level increased, which might at least partially be a result of the recruitment of 4,607 doctors in December 2019 to provide care for the rural population at sub-district hospitals [17]. Higher attendance levels among newly recruited doctors relative to those of existing ones might have contributed to the observed growth in doctors’ attendance at sub-district hospitals.

This study highlights that policymakers in Bangladesh and, given that the results of this study may be generalizable to other low-resource settings, LMICs more generally should undertake major efforts to achieve high attendance levels by HCWs during the COVID-19 pandemic. Several factors likely explain HCWs’ lower work attendance during pandemics – including falling ill themselves, fear of infection [18–20] (which is aggravated by a lack of PPE [4]), and concerns about transmitting the disease to family members [21]. Efforts aimed at addressing these reasons could include ensuring a reliable and sufficient supply of PPE, providing HCWs with comprehensive PPE and emergency preparedness training, following strict isolation protocols at isolation wards, and providing monetary and non-monetary incentives to attend work. In addition, LMICs may choose to implement broader measures to boost human resources for health during the COVID-19 pandemic, such as allowing medical students to graduate early, encouraging retired HCWs to return to work, and mobilizing HCWs from less-affected areas to hardest-hit areas [22,23].

Although this study has several strengths, such as using longitudinal and nation-wide data of fingerprint-verified attendance at all public-sector secondary and tertiary care facilities in a large LMIC to describe how HCWs are responding to the COVID-19 pandemic, it faces several limitations. First, the data only covered the early stages of the COVID-19 pandemic in Bangladesh. Given that the virus will likely continue to spread throughout the country, HCW attendance levels may well decline even further in the coming weeks. Second, the data lacked information on HCW attendance at community clinics and other non-public-sector facilities. However, given that most COVID-19 testing and treatment is provided at secondary and tertiary care facilities (all of which are included in this study), our results pertain to those HCWs who are most heavily involved in the COVID-19 response. Third, the data did not allow us to differentiate between voluntary absence (e.g., due to HCWs’ fear of infection) and absence due to sickness. Finally, we lacked data on attendance across units within hospitals. Since HCWs in certain units – particularly emergency departments and intensive care units – are more heavily involved in caring for COVID-19 patients, analyses stratified by units may provide additional insights to attendance associated with occupational exposures and help inform resource allocation within hospitals.

Low work attendance by HCWs could become a major obstacle to LMICs’ efforts to combat the COVID-19 pandemic. Using Bangladesh as a case study, we show that HCW attendance has been worryingly low in 2019 and 2020 but, encouragingly, has been increasing steadily over time. We also provide evidence that while doctors were more likely to report to duty during the early stages of the COVID-19 pandemic, work attendance by nurses and other staff has dropped since the WHO declared COVID-19 a global emergency. Given that nurses and other staff account for the largest share of the healthcare workforce in LMICs, policymakers should undertake major efforts to boost their attendance levels during the rapidly spreading COVID-19 pandemic.

## Data Availability

The dataset generated and/or analyzed during the current study is publicly available on the Bangladesh Ministry of Health and Family Welfare website [15]: http://103.247.238.92/dghseams/attend/.

## Acknowledgements

None.

## Ethics approval

This study is exempted from ethics approval because the dataset generated and/or analyzed during the current study is publicly available on the Bangladesh’s Ministry of Health and Family Welfare website [15]: http://103.247.238.92/dghseams/attend/.

## Authorship contributions

PG conceived and designed the study. AL and PT extracted the data from the online repository. DD processed the data, conducted the analyses, and visualized the data. DD, PG, and MS interpreted the results. DD wrote the first draft of the manuscript. All authors critically revised the article and approved the final version.

**Appendix Figure 1:**
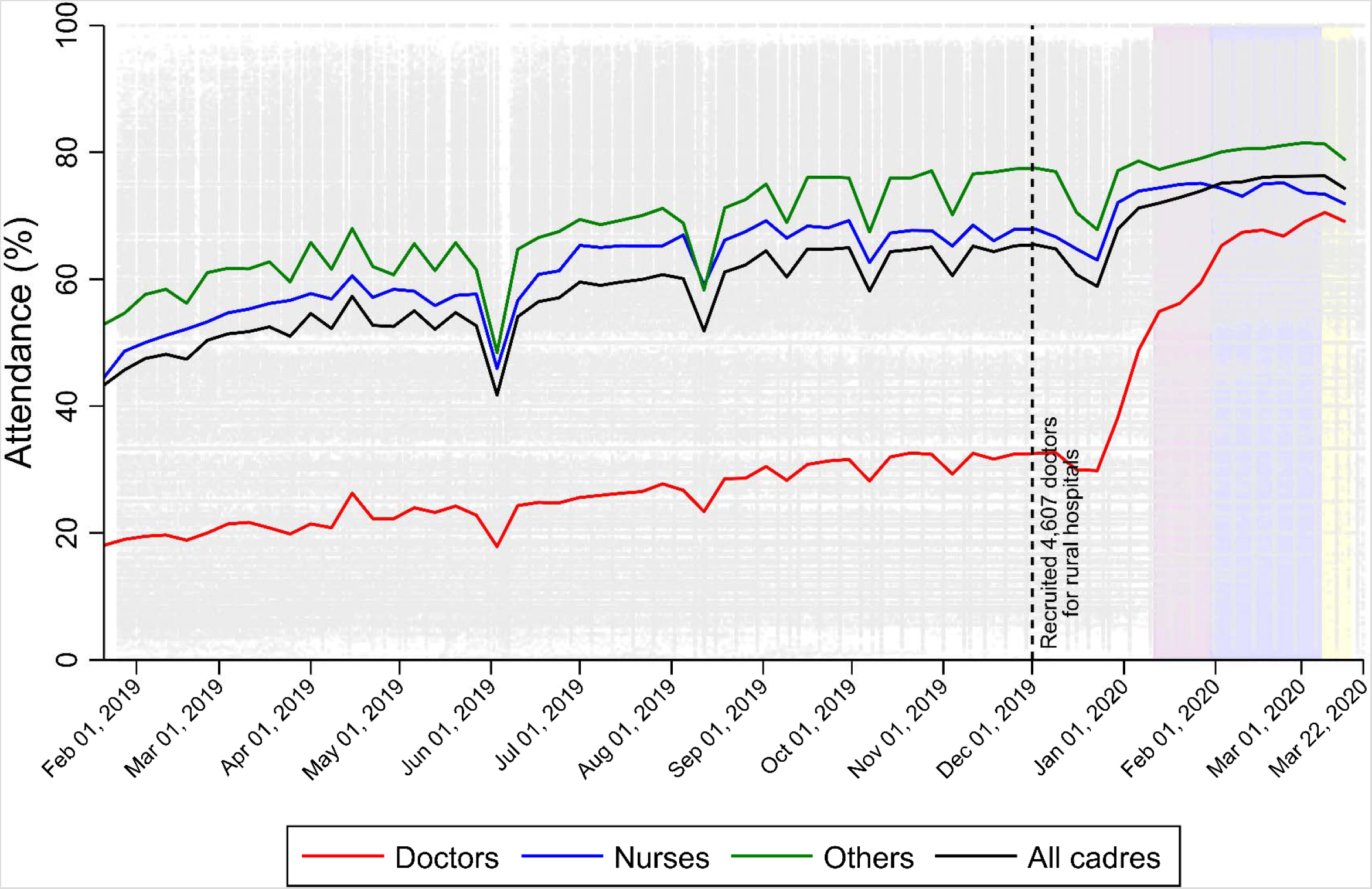
Unsmoothed (Unadjusted) Trends in Weekly Attendance Rates Among Doctors, Nurses, and Other Staff at All Public-Sector Secondary and Tertiary Care Facilities in Bangladesh, by Cadre.

